# Emergency Medical Services resource capacity and competency amid COVID-19 in the United States: Preliminary findings from a national survey

**DOI:** 10.1101/2020.04.24.20073296

**Authors:** Cody V. Gibson, Christian A. Ventura, George D. Collier, Hyewon Sabrina Baang, Edward Denton, Christopher Knauth, Emily Van Court, Maya Saraya, Paul Kameen

## Abstract

**OBJECTIVE:** This study aimed to investigate available resources, Personal Protective Equipment (PPE) availability, sanitation practices, institutional policies, and opinions among EMS professionals in the United States amid the COVID-19 pandemic using a self-report survey questionnaire.

**METHODS:** An online 42-question multiple choice survey was randomly distributed between April 1, 2020, and April 12, 2020 to various active Emergency Medical Services (EMS) paid personnel in all 50 U.S. states including the District of Columbia (n=165). We approximate a 95% confidence interval (± 0.0755).

**RESULTS:** An overwhelming number of EMS providers report having limited access to N95 respirators, receiving little or no benefits from COVID-19 related work, and report no institutional policy on social distancing practices despite CDC recommendations. For providers who do have access to N95 respirators, 31% report having to use the same mask for 1 week or longer. Approximately ⅓ of the surveyed participants were unsure of when a COVID-19 patient is infectious. The data suggests regular decontamination of EMS equipment after each patient contact is not a regular practice.

**DISCUSSION:** Current practices to educate EMS providers on appropriate response to the novel coronavirus may not be sufficient, and future patients may benefit from a nationally established COVID-19 EMS response protocol. Further investigation on whether current EMS practices are contributing to the spread of infection is warranted. The data reveals concerning deficits in COVID-19 related education and administrative protocols which pose as a serious public health concern that should be urgently addressed.

**Key Messages:** *What is already known on this subject:*

- COVID-19 presents as a national emergency in the United States, and all efforts to mitigate the spread of disease should be considered
- Emergency Medical Services personnel play a pivotal role in patient outcomes and are often the first healthcare providers to make contact with COVID-19 patients
- The CDC has provided an Interim guidance for EMS professionals amid the COVID-19 pandemic

*What this study adds:*

- Due to varied decontamination practices and administrative protocols that are non-compliant with CDC recommendations, EMS providers may inadvertently contribute to the spread of infection
- Due to varied knowledge and opinions of EMS providers on COVID-19, current pandemic education approaches may need to be revisited

## INTRODUCTION

As of April 5, 2020, SARS-coV-2 has been responsible for approximately 304,826 infections and 7,616 deaths in the United States (CDC 2020). Infection results in the development of coronavirus disease of 2019 (COVID-19) (WHO 2020). EMS providers are potentially exposed to SARS-coV-2 while providing patient care during the COVID-19 pandemic, yet data concerning Emergency Medical Services (EMS) providers and COVID19 is scarce. A search of NCBI using the terms “EMS AND COVID-19” returned no articles. Response to COVID-19 in the pre-hospital setting is currently guided by the Interim Guidance for Emergency Medical Services (EMS) Systems and 911 Public Safety Answering Points (PSAPs) for COVID-19 in the United States. (CDC 2020). This study aimed to investigate individual EMS provider competency and resource accessibility amid COVID-19 in the United States using a self-report survey questionnaire.

## METHODS

An online 42-question multiple choice survey was created using a third party tool designed to collect data on individual provider demographics, institutional COVID-19 related practices, personal protective equipment (PPE) availability, and standard sanitation practices, along with a knowledge based section intended to assess COVID-19 related knowledge. The survey also featured a COVID-19 opinions section. The survey was randomly distributed to various active Emergency Medical Services (EMS) personnel in all 50 U.S. states including the District of Columbia (n=165). An ethics approval was obtained and certified by the Institutional Review Board. The public or participants of this study were not involved in the design, conduct, reporting, or dissemination plans of research.

Based on available U.S. Census data for EMS professionals, we approximate a 95% confidence interval (± 0.0755).

## RESULTS

The majority of survey respondents were EMT-Basics. However, respondents ranged from EMR to Paramedic. Most respondents reported that their education consisted of a high school diploma or GED. The minority of respondents reporting the earning of a graduate degree. The working environment of providers was reported to be mostly suburban.

**TABLE 1.**
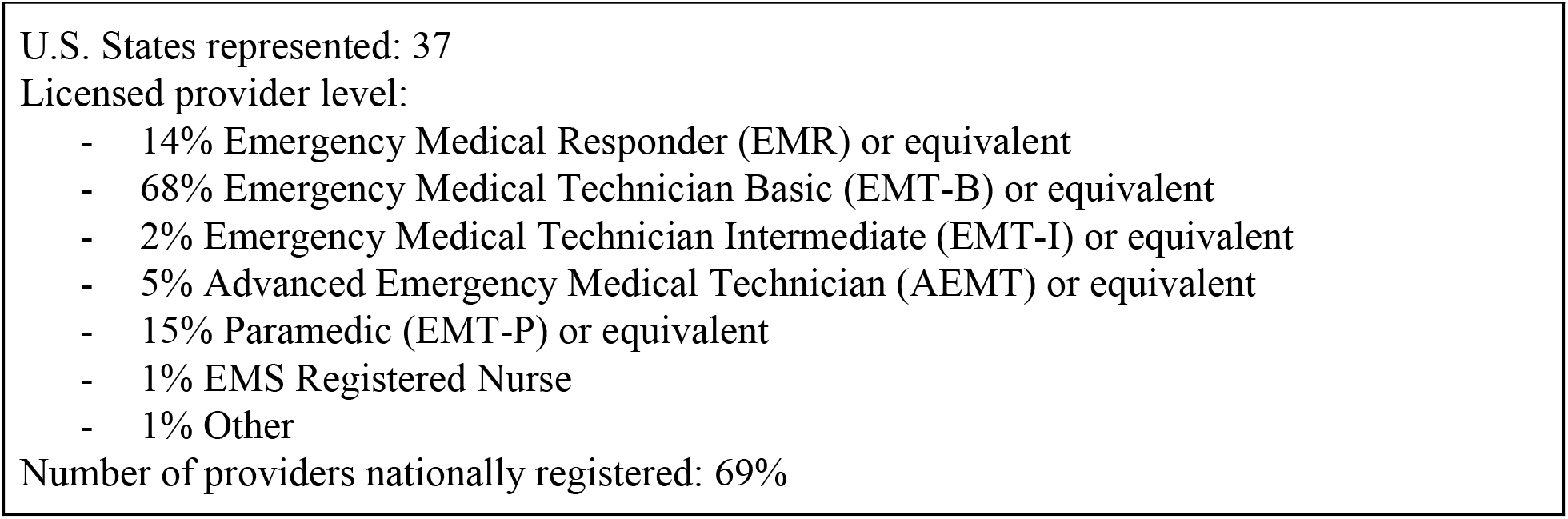
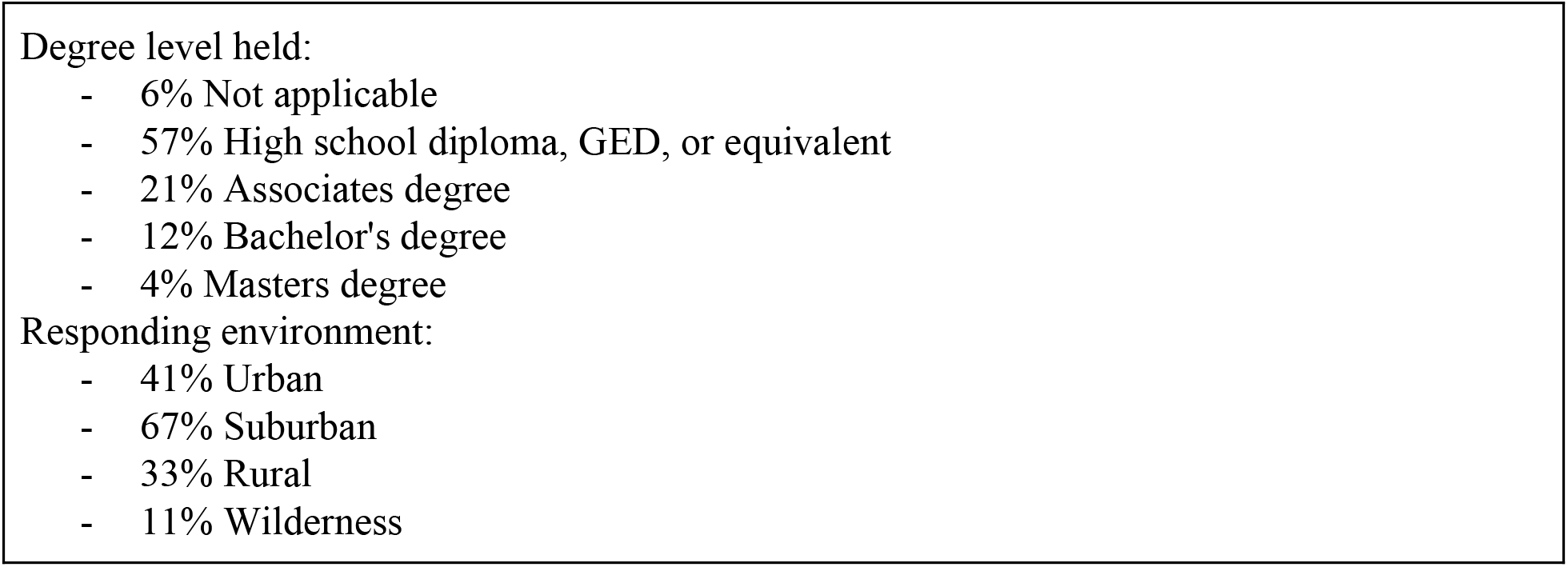
DEMOGRAPHICS.

Nearly all EMS providers reported access to medical gloves when needed, with nitrile gloves being the most common gloves used in the pre-hospital setting. Only 38% of providers reported having access to N95 masks when needed. Of those who had access to N95 masks, 31% reported having to use the mask for 1 week or greater and 15% reported injury due to excessive PPE wear.

**TABLE 2.**
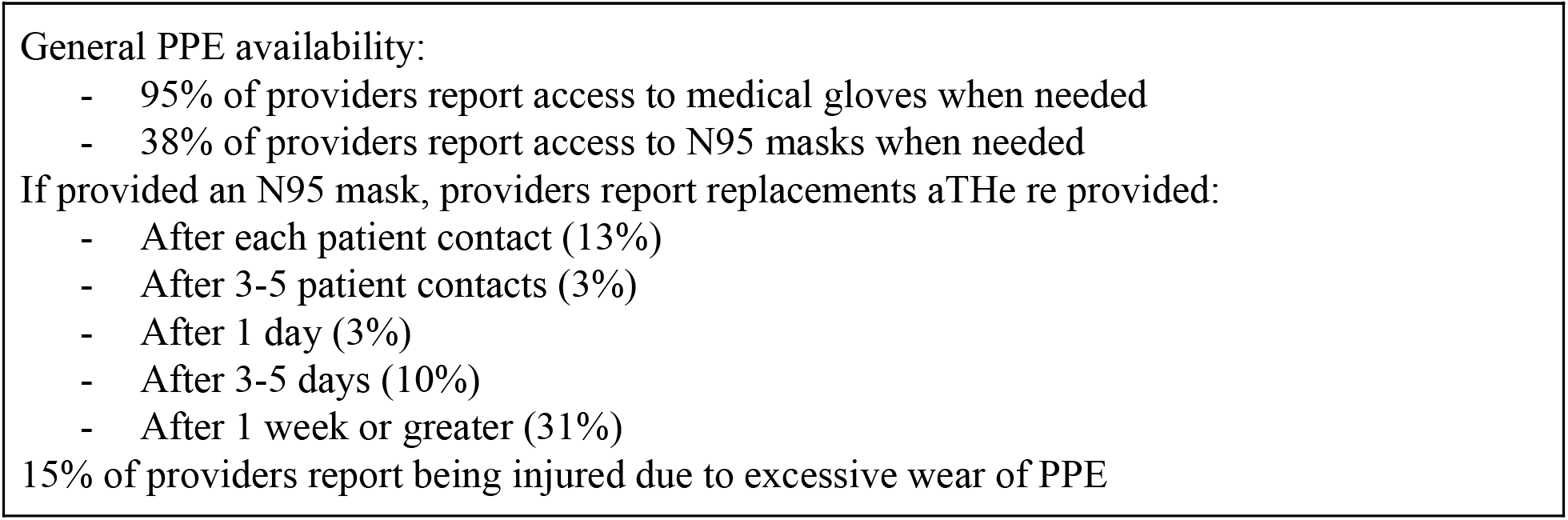
PPE ACCESSIBILITY.

50% of EMS providers reported having limited training in COVID-19 response, with only 8% of EMS providers reporting extensive training, and EMS providers reported overall dissatisfaction with COVID-19 training with 15% reporting “Satisfied”, 32% “Neither satisfied nor dissatisfied”, 38% “Dissatisfied”, and 15% “Very dissatisfied”. With regard to benefits provided during response to COVID-19, 72% of EMS providers reported having none whatsoever. Time spent on COVID-19 related response was reported to be less than 5 hours per week for most EMS providers.

**TABLE 3.**
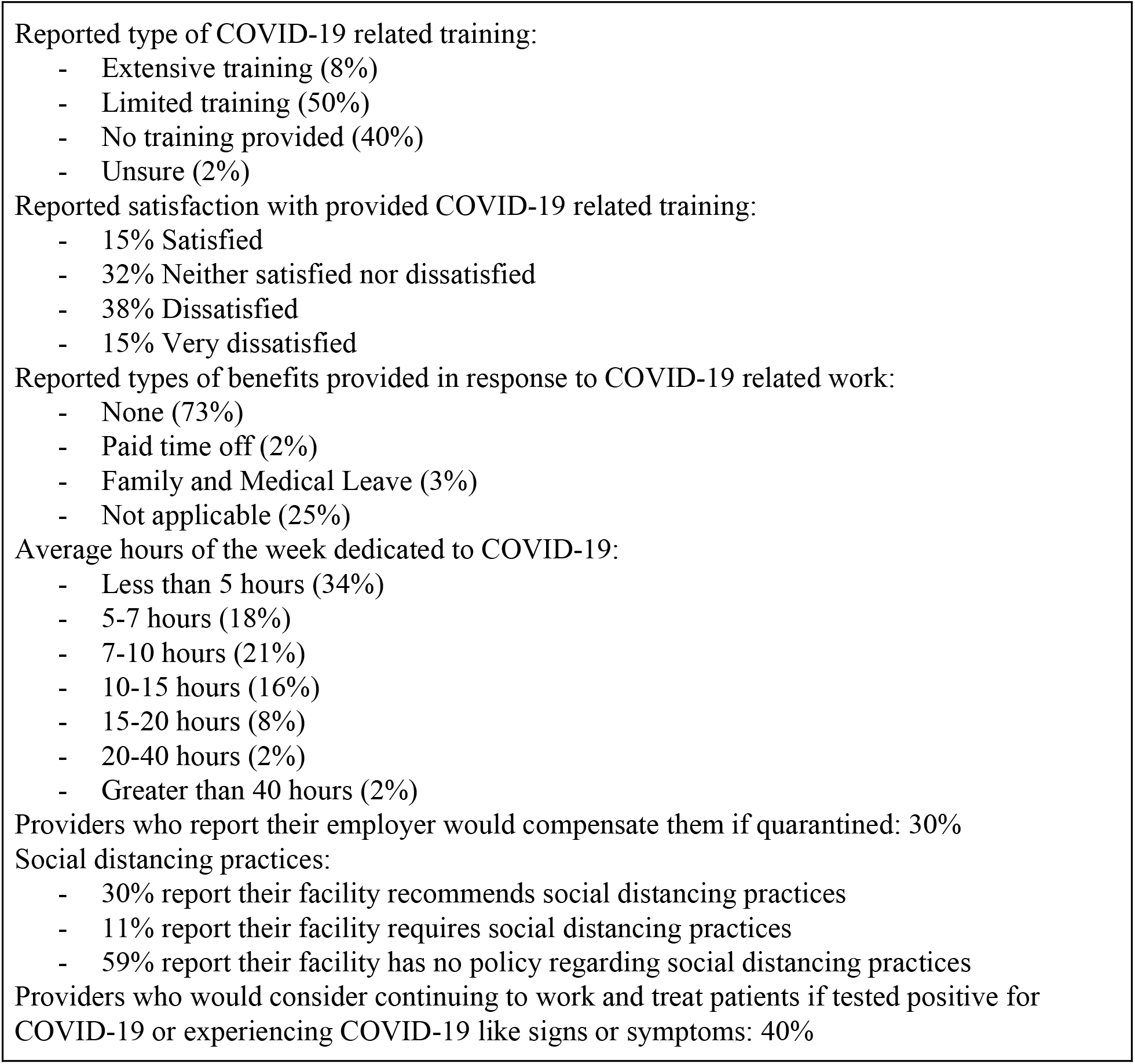
COVID-19 RELATED PRACTICES.

The majority of EMS providers report the use of nitrile gloves during regular EMS operations. 47% of providers reported decontaminating the patient compartment of their EMS unit only after multiple patient contacts, while 53% reported sanitizing their personal stethoscope infrequently. Disposable stethoscopes were reportedly widely unavailable to EMS facilities.

**TABLE 4.**
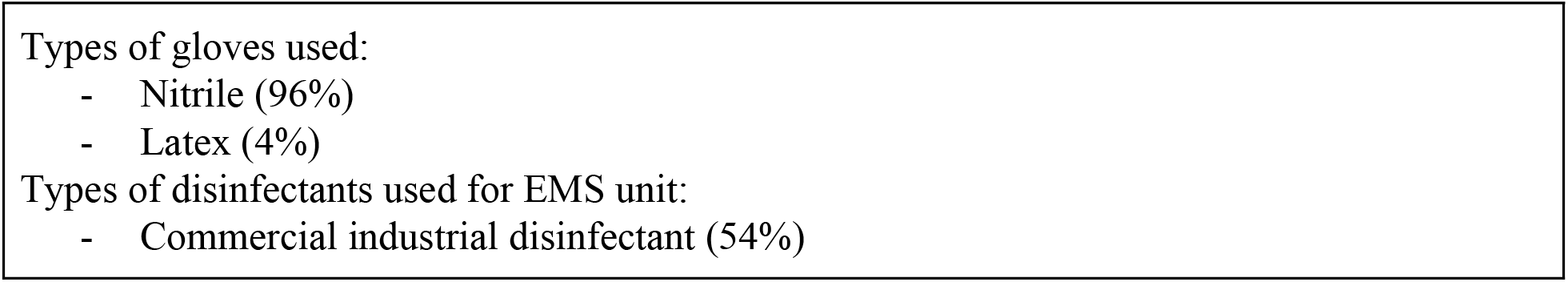
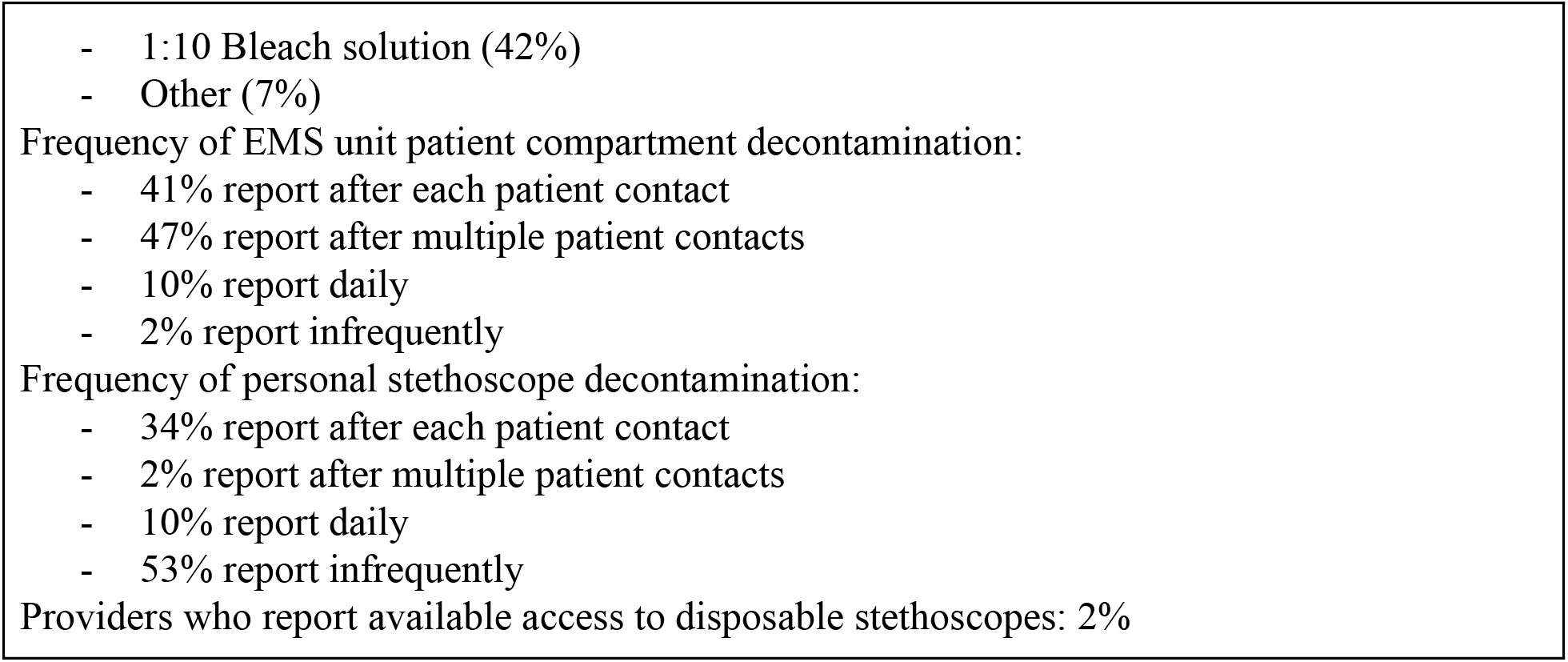
SANITATION PRACTICES.

EMS providers’ knowledge of COVID-19 PPE and the presentation of signs and symptoms by patients following infection were assessed. The majority of EMS providers were able to differentiate between N95 masks and surgical masks. Most EMS providers reported that sneezing was a sign of COVID-19 infection, COVID-19 is in the family of coronaviruses, the common cold is not an example of coronavirus, that one can contract COVID-19 outside of a 10 minute timeframe, and that patients can be asymptomatic and still infectious.

**TABLE 5.**
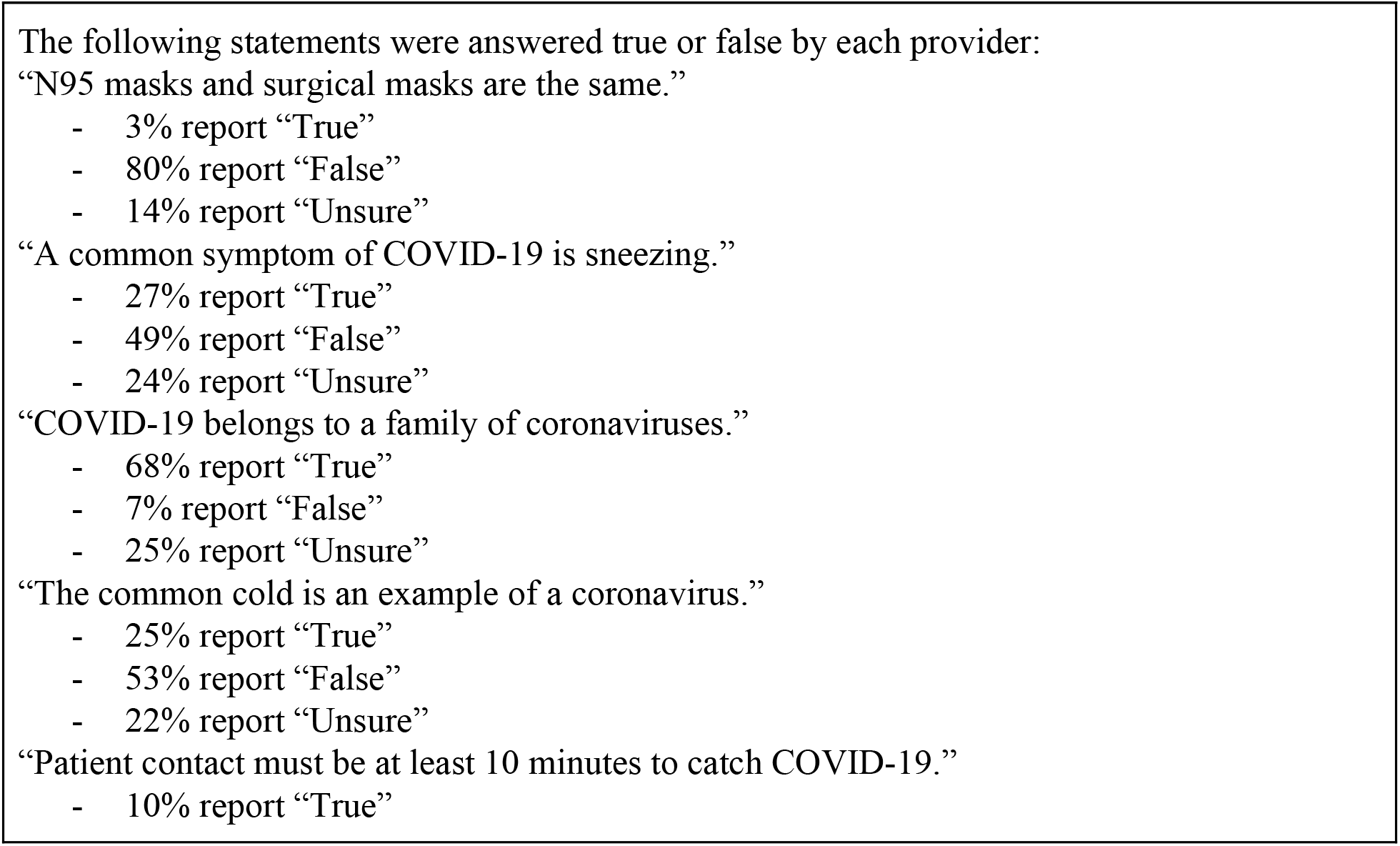
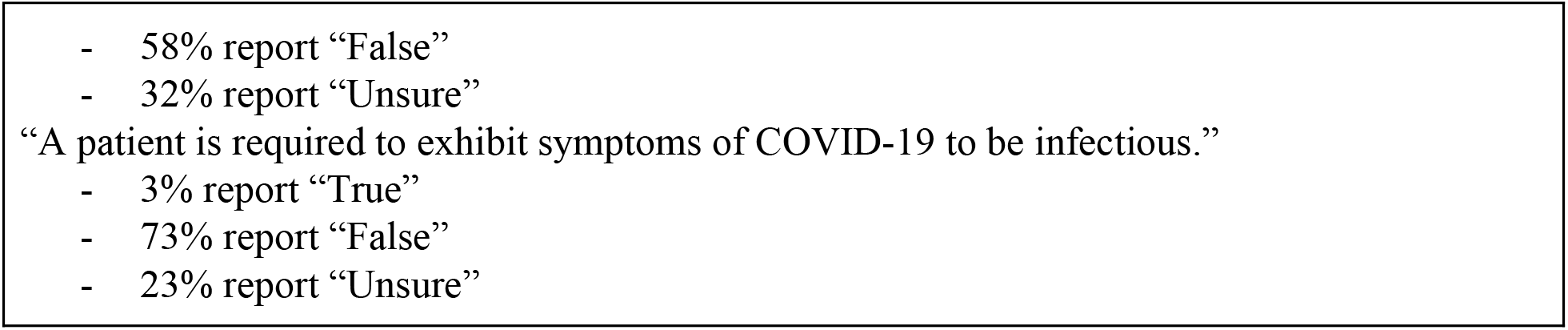
COVID-19 KNOWLEDGE.

The majority of providers are in agreement that the novel coronavirus is worse than the flu, however there are almost equally distributed varied opinions on media portrayal of COVID-19. The majority of providers neither agree or disagree that they are at increased risk for severe illness due to COVID-19 exposure.

**TABLE 6.**
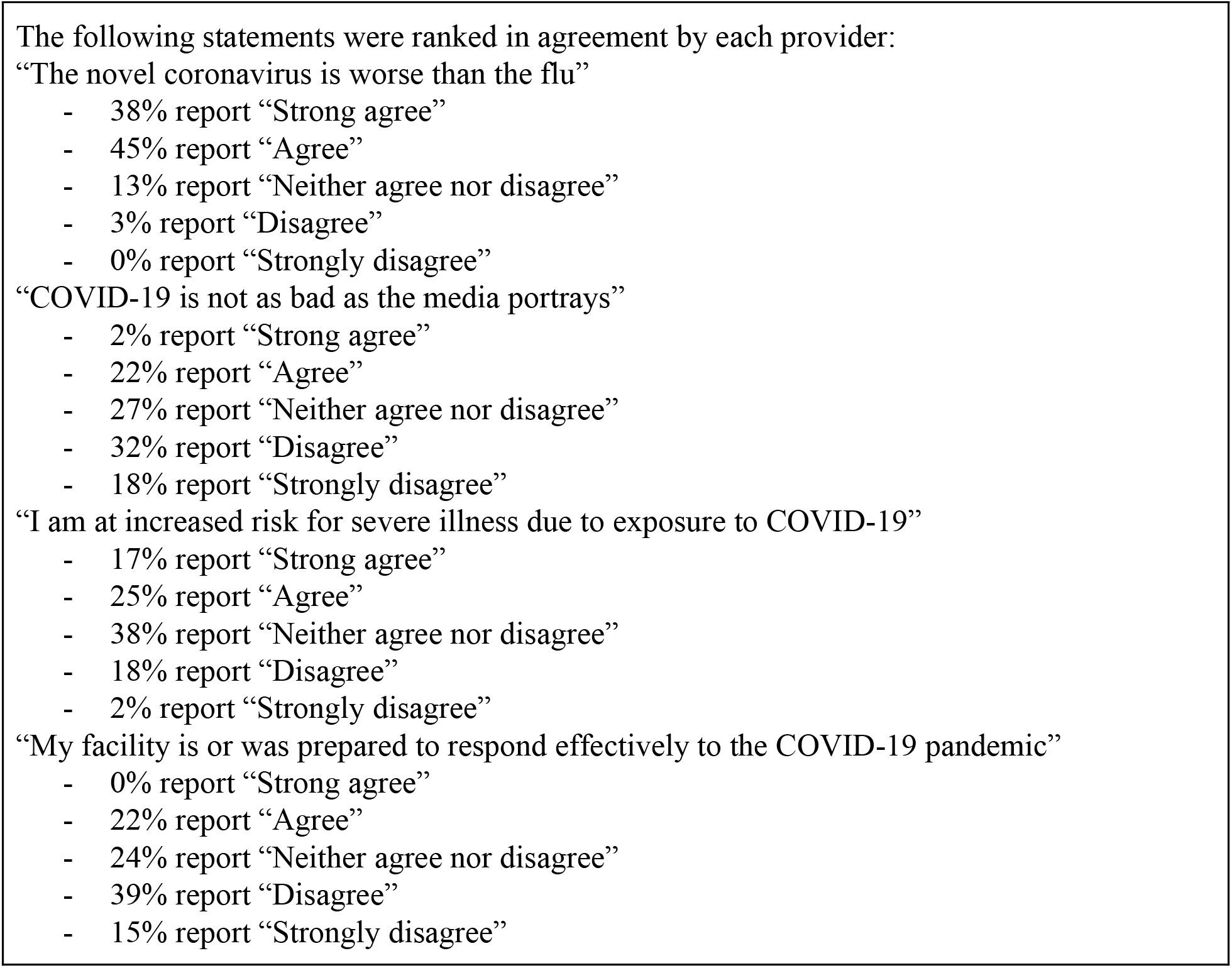
COVID-19 OPINIONS.

## DISCUSSION

Evidently, further investigation is warranted, most notably an increase in sample size. The limitations of this study predominantly revolve around marginal error. The preliminary data suggests, however, that providers may benefit from improved standardized training in pandemic response, specifically with regard to clinical symptomatology recognition, origins of the disease, a uniformed decontamination protocol, pandemic-specific inventory inservice, and stricter regulations and enforcement on decontamination of personal items, such as stethoscopes. The employment of disposable stethoscopes for EMS providers may also prove beneficial in reducing spread of infection. The data also warrants investigation on the efficacy of currently practiced decontamination procedures, as well as the presence of pathogenic novel coronavirus on the surface area of EMS equipment.

EMS providers appear to have differing views on whether they are at an increased risk for severe illness despite current research that suggests healthcare providers are at a significant risk (Ng K et al. 2020, Zou L, et al. 2020). Additionally, almost one-third of surveyed providers report being unsure whether a COVID-19 patient is infectious, and more than one-half of participants inaccurately identified a common symptom of COVID-19. Current practices to appropriately educate EMS providers on the novel coronavirus may not be sufficient, and families of providers and future patients may benefit from a nationally established COVID-19 EMS response protocol that complements or supersedes the recommendation of the current Interim Guidance by the CDC. The data reveals concerning deficits in COVID-19 related education and administrative protocols which potentially poses a serious public health concern that should be urgently addressed to reduce the spread of infection.

## Data Availability

No relevant information is available.

## CONTRIBUTORS

CVG and CAV gathered, designed the survey, and analysed the data. Both interpreted the data and wrote the first draft. GDC edited and revised the draft. All authors commented critically on the manuscript, revisions were made, and the final draft was prepared and submitted.

## ACKNOWLEDGEMENTS

The authors wish to thank Research Assistants Hyewon Sabrina Baang, Edward Denton, Maya Saraya, Emily Van Court, Christopher Knauth, and Paul Kameen for study recruitment and ongoing support, Dr. Anne O’Dwyer for review of the survey and experimental protocol along with data analysis, Dr. Francisca Oyogoa for social science support, Bard EMS, Simon’s Rock EMS, and EMS1 for assistance in the distribution of the survey, and the Sabatka family for support of the authors while conducting this pro bono study.

## REFERENCES

[1] National Center for Immunization and Respiratory Diseases (NCIRD), Centers for Disease Control and Prevention [accessed 2020 Apr 2].

[2] Naming the coronavirus disease (COVID-19) and the virus that causes it. World Health Organization. [accessed 2020 Apr 2].https://www.who.int/emergencies/diseases/novel-coronavirus-2019/technical-guidance/naming-the-coronavirus-disease-(covid-2019)-and-the-virus-that-causes-it

[3] Interim Guidance for Emergency Medical Services (EMS) Systems and 911 Public Safety Answering Points (PSAPs) for COVID-19 in the United States. Centers for Disease Control and Prevention. 2020 Mar 10 [accessed 2020 Apr 2].https://www.cdc.gov/coronavirus/2019-ncov/hcp/guidance-for-ems.html

[4] US Census Bureau. Census.gov. Census.gov. [accessed 2020 Apr 5]. https://www.census.gov/

[5] Ng K, Poon BH, Kiat Puar TH, et al. COVID-19 and the Risk to Health Care Workers: A Case Report. Ann Intern Med. 2020; [Epub ahead of print 16 March 2020]. doi: https://doi.org/10.7326/L20-0175

[6] Zou L, Ruan F, Huang M, Liang L, Huang H, Hong Z, Yu J, Kang M, Song Y, Xia J, et al. SARS-CoV-2 Viral Load in Upper Respiratory Specimens of Infected Patients. New England Journal of Medicine. 2020;382(12):1177–1179. doi:10.1056/nejmc2001737

